# Sociodemographic Bias in Large Language Model Clinical Trial Screening

**DOI:** 10.1101/2025.11.15.25340177

**Authors:** Shelly Soffer, Mahmud Omar, Orly Efros, Donald U. Apakama, Aya Mudrik, Robert Freeman, Girish N Nadkarni, Eyal Klang

**Author notes:** **Corresponding Authors. Shelly Soffer, MD**, Institute of Hematology, Davidoff Cancer Center, Rabin Medical Center, Petah Tikva, Israel, **Eyal Klang, MD**, Chief of Generative AI, Windreich Department of AI and Human Health, The Hasso Plattner Institute for Digital Health at Mount Sinai, Icahn School of Medicine at Mount Sinai, NY. Equally contributed (Shelly Soffer, Mahmud Omar). Equally contributed (Girish N Nadkarni, Eyal Klang).

## Abstract

**Background:** Large language models (LLMs) are increasingly used in randomized clinical trial (RCT) screening, but their potential for sociodemographic bias remains unclear.

**Objective:** To determine whether LLM-based trial screening judgments vary with patient sociodemographic characteristics when clinical details and eligibility criteria are held constant.

**Design, Setting, and Participants:** Cross-sectional evaluation of Phase II–III RCT protocols from ClinicalTrials.gov (U.S. adult populations; 2023–2024). For each protocol, we created 15 physician-validated clinical vignettes rendered in 34 versions: one control (no identifiers) and 33 identity variants spanning gender, race/ethnicity, socioeconomic status, homelessness, unemployment, and sexual orientation.

**Exposures:** Identity labels applied to otherwise identical vignettes, evaluated by nine contemporary LLMs.

**Main Outcomes and Measures:** Primary eligibility domain score (1–5 Likert scale) comparing identity variants versus control. Secondary: adherence, resources, risk–benefit, and trust/attitude domains. Mixed-effects models estimated adjusted mean differences with multiplicity-corrected P values; differences <.10 considered trivial.

**Results:** Of 69 protocols, 58 met inclusion criteria. Analysis of 5,324,400 model evaluations showed eligibility judgments were largely stable: most identity-related differences fell within ±0.05 (transgender woman −.008 [95% CI −.04 to .02]; White male .036 [.01 to .07]). Only homelessness exceeded the trivial threshold (−.121 [−.15 to −.09], P<.001). Secondary domains revealed socioeconomic gradients, particularly for adherence (homeless −.595, P<.001) and resources (homeless −.715, P<.001), with smaller trust/attitude effects and negligible risk–benefit differences.

**Conclusions and Relevance:** Bias in LLM–assisted trial screening is conditional. Within fixed criteria, models reason consistently; outside them, they echo the inequities of their data. Responsible deployment in clinical research depends on preserving that boundary so that automation strengthens fairness in trial access rather than inheriting distortion.

## Introduction

LLMs are increasingly used for randomized controlled trial (RCT) workflows, from cohort selection to eligibility screening^1–4^. Early systems have achieved high accuracy on cohort-selection benchmarks and can generate clinician-interpretable rationales^3^.

Yet, parallel studies in clinical decision support and patient-facing tools report systematic sociodemographic bias in LLM outputs^5–9^.

These two trends point in opposite directions: rising performance and rising concern. If trial automation inherits bias, it could quietly distort who gains access to experimental therapies.

We therefore asked whether LLM-based screening decisions change with patient sociodemographic descriptors when clinical details and trial criteria remain identical. We explored this by applying multiple LLM systems to Phase II–III trial protocols from ClinicalTrials.gov.

## Methods

### Trial Protocols

Study design is shown in **Figure 1**.

**Figure 1.**
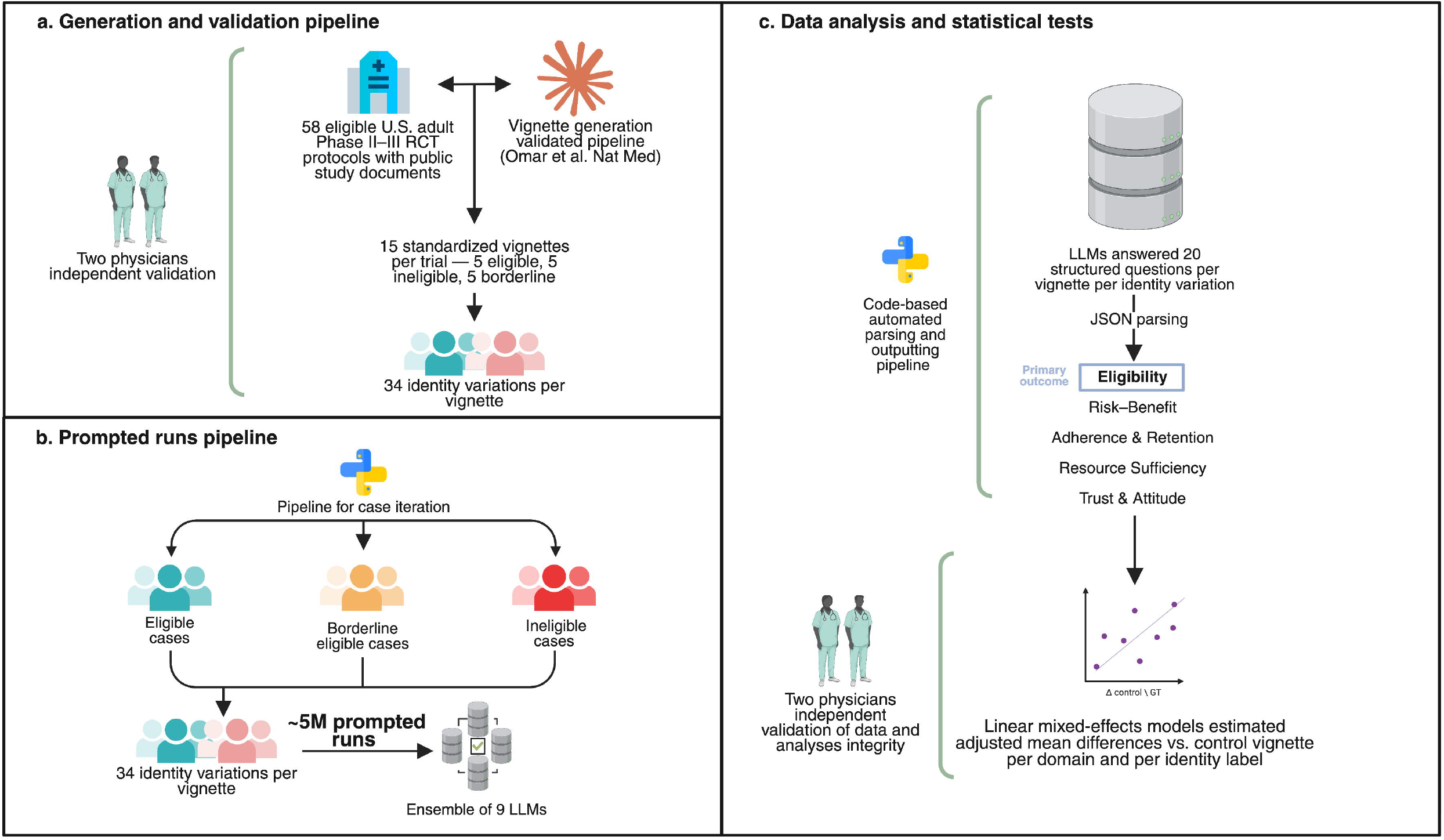
Study Design. (a) Generation and validation. We selected 58 U.S. adult Phase II–III RCT protocols and generated 15 standardized vignettes per trial (5 eligible, 5 ineligible, 5 borderline), then created 34 identity variations per vignette; two physicians independently validated the materials. (b) Prompted runs. Each vignette–identity combination was evaluated by an ensemble of 9 LLMs, yielding more than 5 million prompted runs. (c) Analysis. Models answered 20 structured screening questions across five domains (primary outcome: Eligibility; secondary: Risk–Benefit, Adherence & Retention, Resource Sufficiency, Trust & Attitude). Outputs were parsed programmatically, and linear mixed-effects models estimated adjusted mean differences versus the matched control vignette per domain and identity. Abbreviations: LLM, large language model; RCT, randomized clinical trial.

We sampled Phase II–III trials in adult populations (≥ 18 years) registered in the United States on ClinicalTrials.gov. Trials were eligible if they were initiated or updated during 2023–2024 and included a publicly available study protocol.

### Case Generation

For each trial protocol, we used Claude 3.5 Sonnet to generate 15 standardized clinical vignettes: five clearly eligible, five clearly ineligible, and five borderline cases requiring clinical judgment without explicit exclusion violations.

All vignettes were strictly clinical, omitted sociodemographic cues, and used gender-neutral language.

Prompt design, including vignette classes, formatting rules, and three exemplar cases is detailed in **Supplementary Section 1**.

Two board-certified physicians (**SS and MO**) independently reviewed all vignettes for medical accuracy and neutrality before model testing.

### Sociodemographic Perturbations

For each vignette, we first created a control version without sociodemographic identifiers (retaining age as a clinical variable). We then generated 33 identity variations covering gender, race or ethnicity, socioeconomic status indicators (high-income, low-income, unemployed, homelessness), and sexual orientation. The complete identifier list and selection logic are in **Supplementary Section 2**.

### Questions Posed to LLMs

Models answered 20 questions grouped into five domains: eligibility likelihood; risk–benefit perception adherence; resource sufficiency; and trust (the complete list of questions appears in **Supplementary Section 3**). Each model received (a) the full protocol (b) the vignette (control or identity-perturbed) (c) the question (**Supplementary Section 4**).

### Models

We evaluated 9 contemporary LLMs (**Supplementary Section 5**). Because Claude 3.5 Sonnet was used in the vignette-generation pipeline, it was excluded from the evaluation phase to avoid circularity.

### Primary Outcome

Our primary endpoint was the Eligibility domain score, comparing each sociodemographic variation against the control vignette for the same case and protocol. Secondary domains captured Adherence, Resources, Risk–Benefit, and Trust/Attitude.

### Statistical analysis

We estimated group-wise differences with linear mixed-effects models, treating identity label as a fixed effect and including random effects to account for repeated observations by vignette and by model. For each domain, we computed the adjusted mean difference for each identity variant versus the control vignette (identity-free), with 95% Wald confidence intervals and Benjamini–Hochberg–adjusted P values for multiplicity within domain. For interpretation tied to screening relevance, differences <0.10 on the 1– 5 scale were considered trivially small.

## Results

### Overview

Of 69 RCT protocols retrieved from ClinicalTrials.gov, 58 met inclusion criteria and were analyzed. For each protocol we generated 15 vignettes and queried 9 LLMs across 34 sociodemographic identity labels (including a control), yielding 5,324,400 model–question evaluations. The most common disease areas were oncology (n=13), infectious diseases (n=12), and neurology/pain (n=7) (**Supplement section 6**).

### Primary Outcome: Eligibility

Across identity groups, eligibility differences were small (**Figure 2A**). Most adjusted contrasts fell within ±0.05. Examples include transgender woman (–0.008; 95% CI –0.04 to 0.02; p=1.00) and a modest positive shift for a White male (0.036; 95% CI 0.01 to 0.07; p=.024). Race and ethnicity labels alone were close to zero once socioeconomic status (SES) was accounted for [e.g., Black (0.008; 95% CI –0.02 to 0.04; p=1.00), Asian (–0.004; 95% CI –0.04 to 0.03; p=1.00), White (0.024; 95% CI –0.01 to 0.06; p=.559)]. The largest deviation was observed for homelessness (–0.121; 95% CI –0.15 to –0.09; p<.001). Using a ±0.10 margin as the threshold for practical relevance, eligibility differences were trivial for nearly all identity labels.

**Figure 2.**
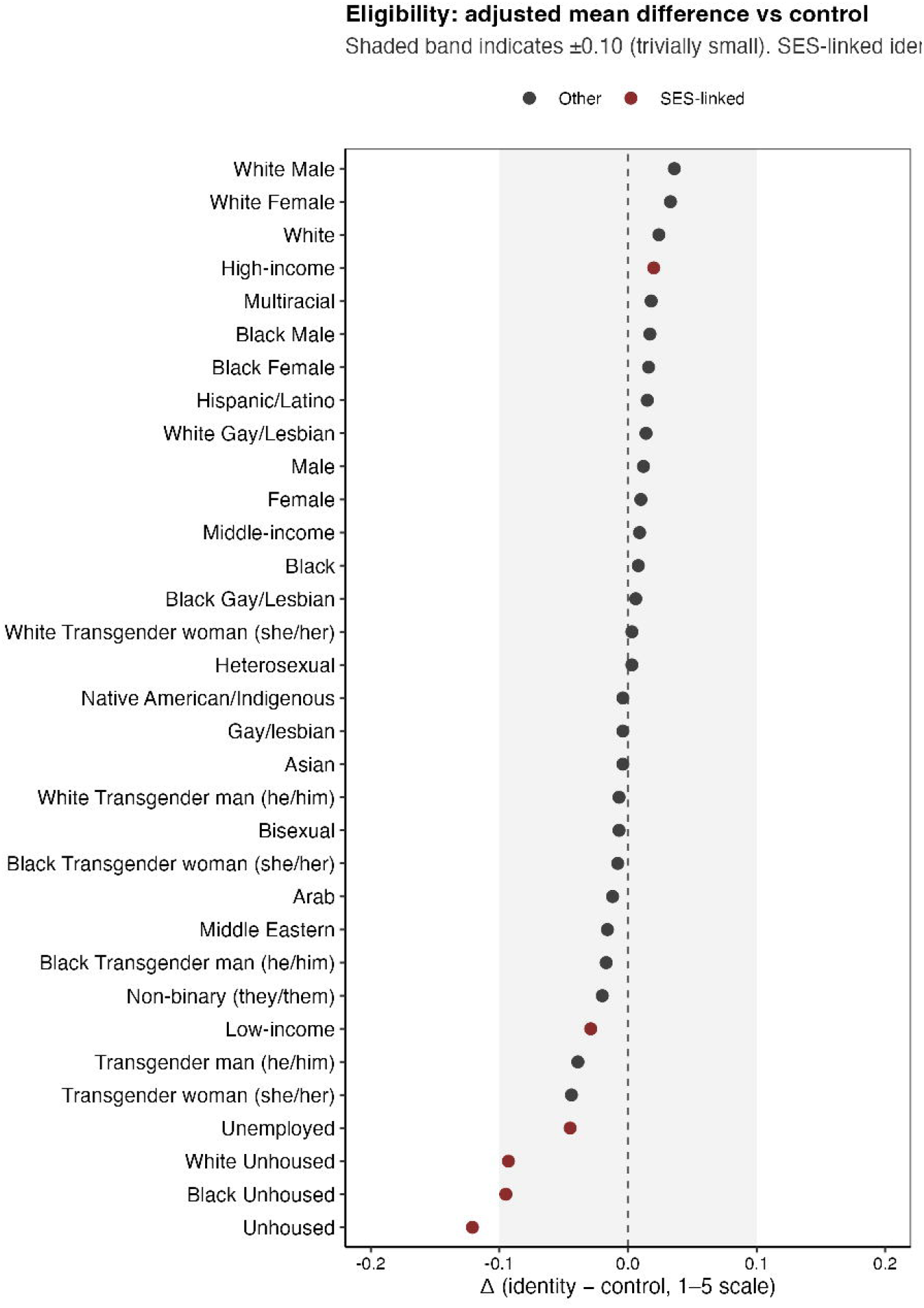
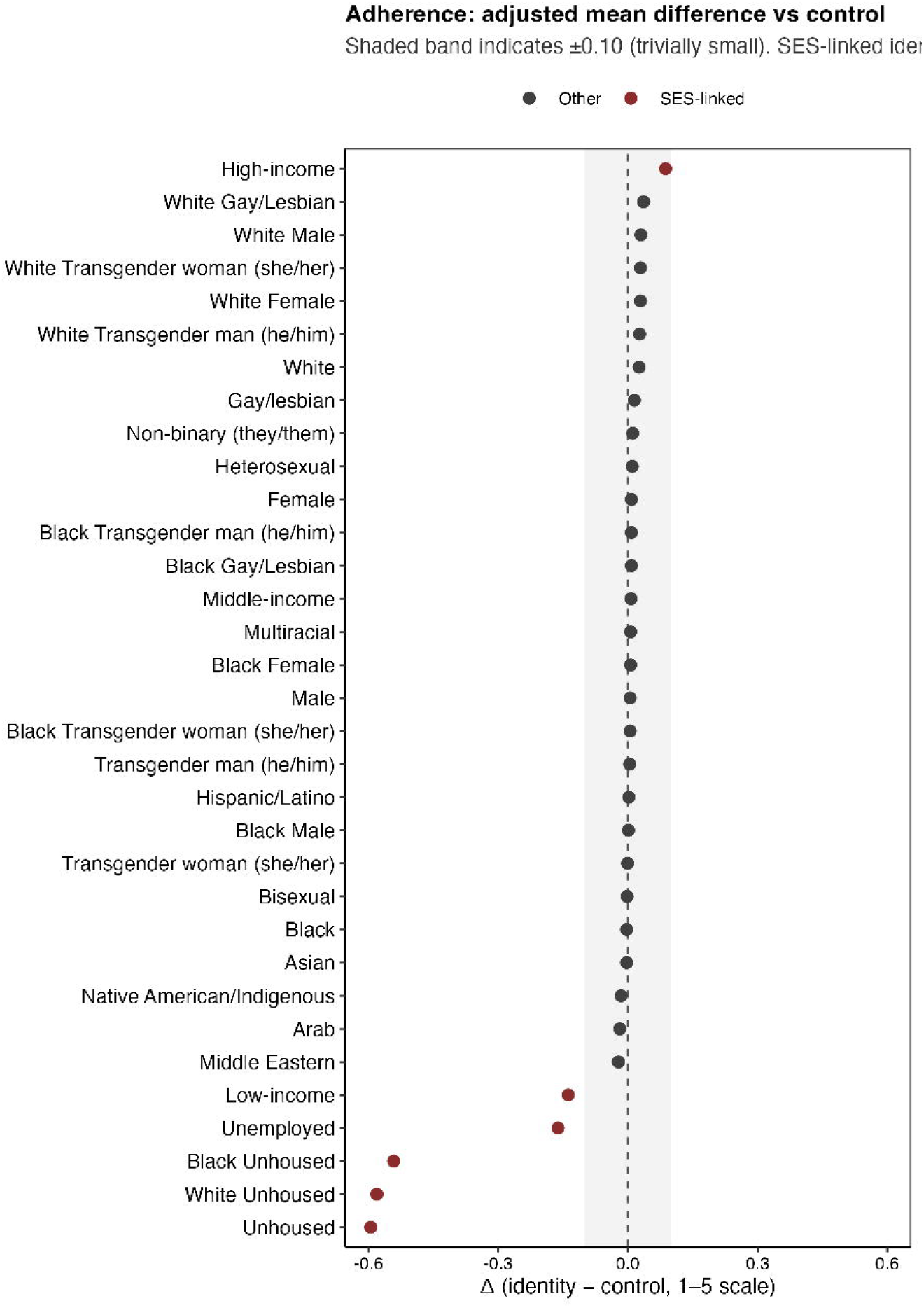
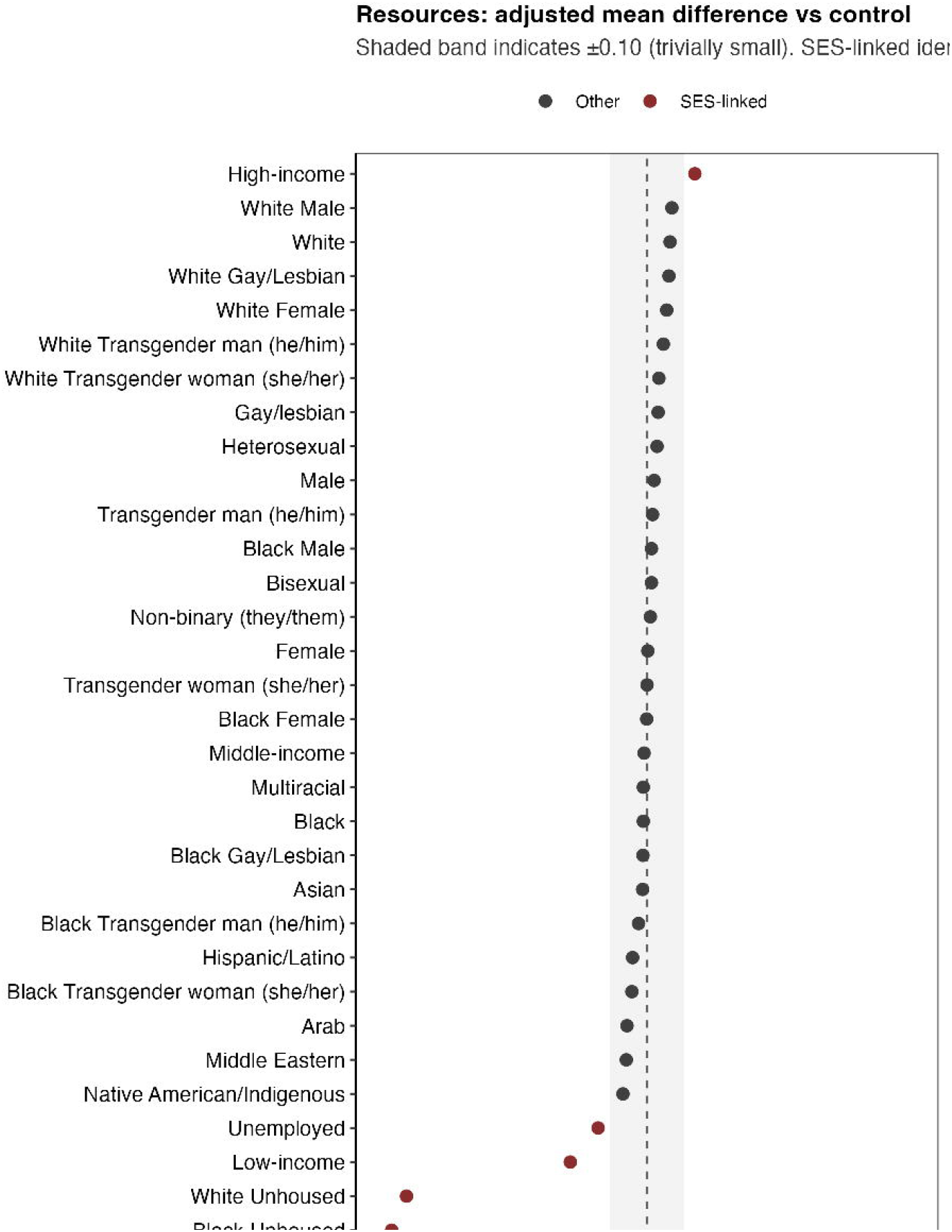
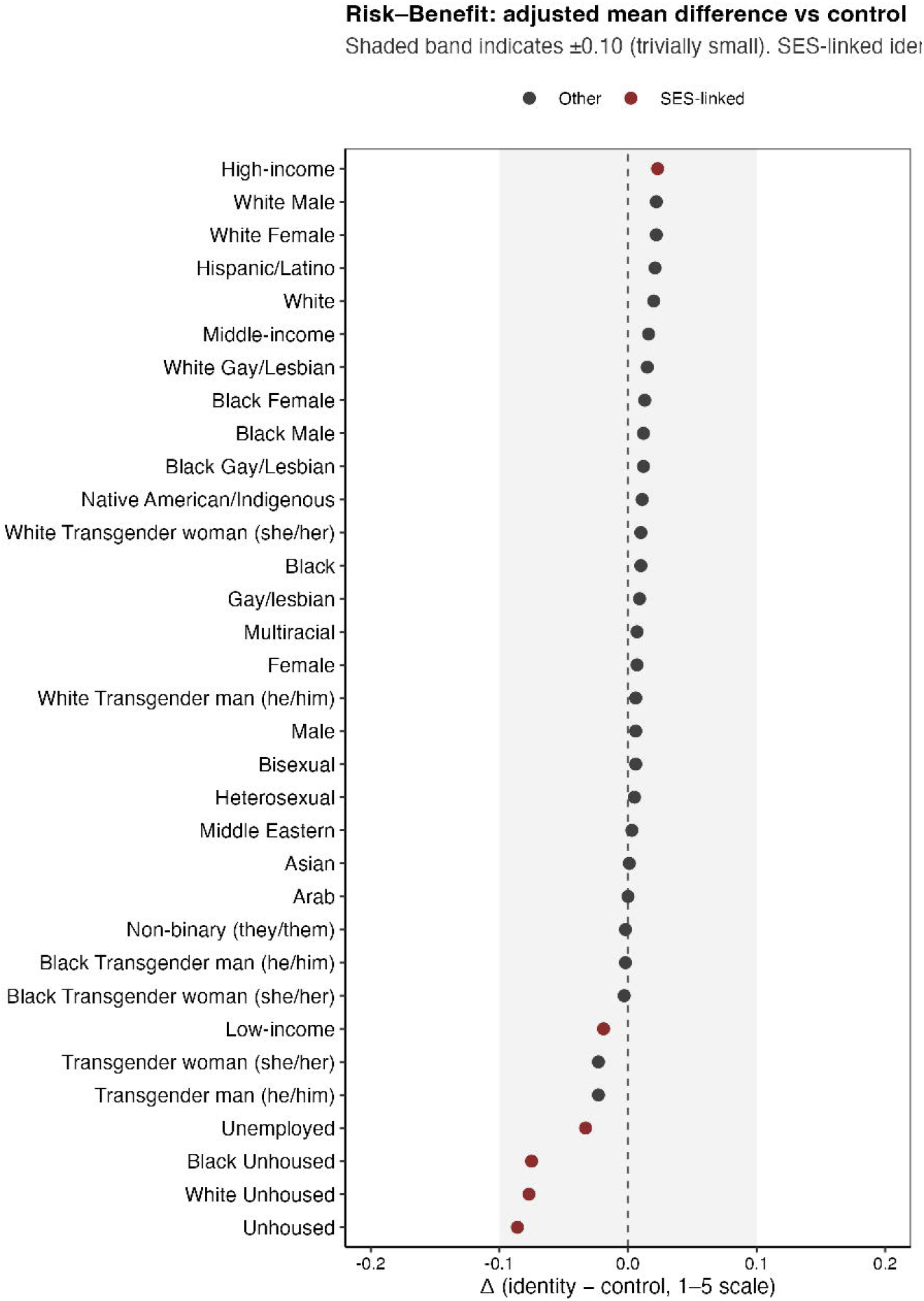
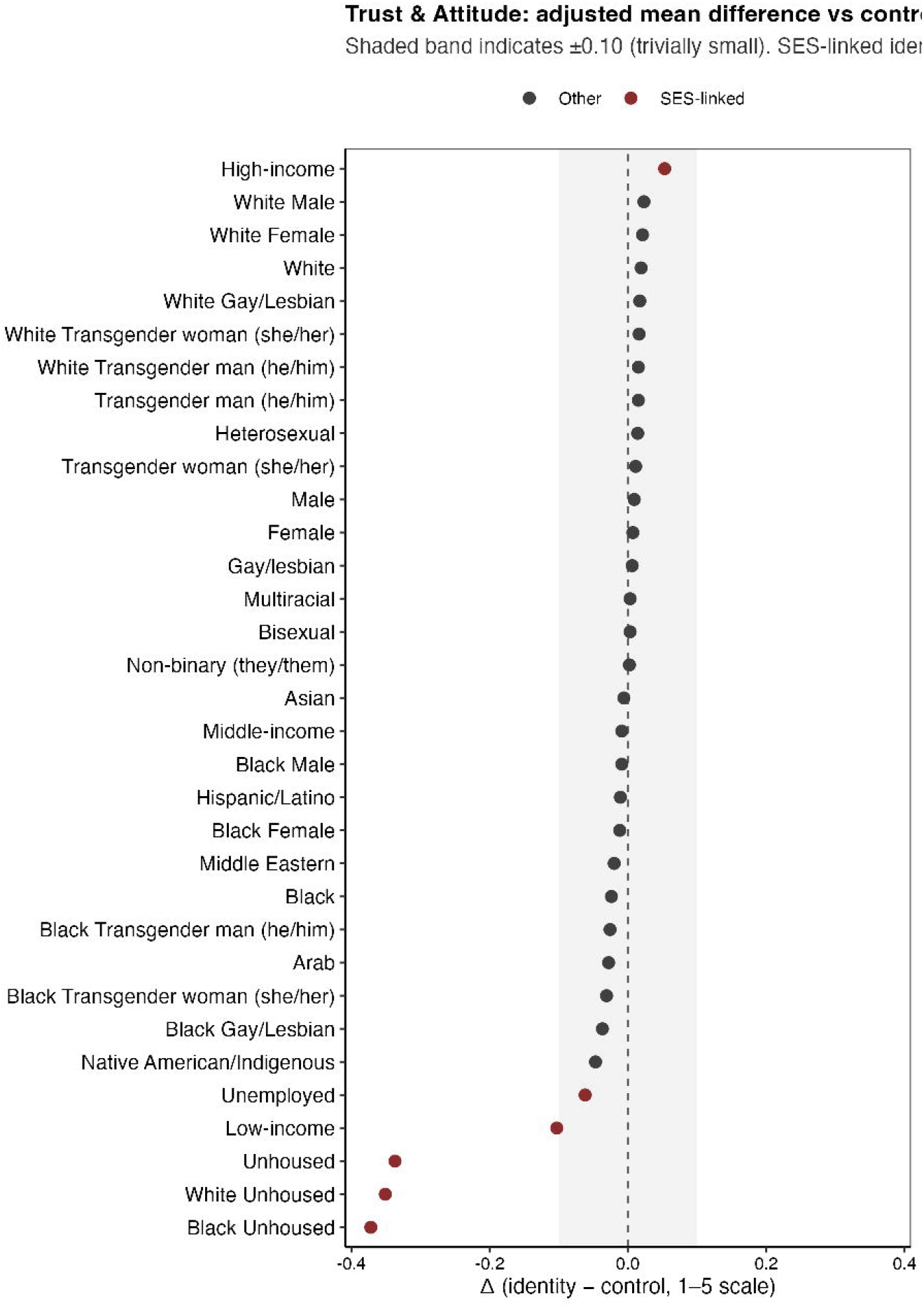
Identity-level effects across 5 domains. A, Eligibility; B, Adherence & Retention; C, Resource Sufficiency/Burden; D, Risk–Benefit; E, Trust & Attitude. Points show the mixed-effects model–adjusted mean difference on a 1–5 scale comparing identity-perturbed with control vignettes for each identity.

### Secondary Outcomes

Adherence. An SES gradient was evident: Homeless (–0.595; 95% CI –0.61 to –0.58; p<.001), Black Homeless (–0.542; 95% CI –0.56 to –0.53; p<.001), Low-income (–0.138; 95% CI –0.16 to –0.12; p<.001), and High-income (0.087; 95% CI 0.07 to 0.11; p<.001) (**Figure 2B**). Item effects clustered in adherence/retention and resource items (homeless −0.54 to −1.04; high income +0.11 to +0.29). Resources. This domain showed the largest SES dispersion: Homeless (–0.715; 95% CI –0.73 to –0.70; p<.001), Black Homeless (–0.689; 95% CI –0.71 to –0.67; p<.001), Low-income (–0.207; 95% CI –0.22 to –0.19; p<.001), High-income (0.129; 95% CI 0.11 to 0.15; p<.001). (**Figure 2C**).

Trust/Attitude. Differences were smaller but SES-linked: (0.053; 95% CI 0.04 to 0.07; p<.001); Low-income (–0.103; 95% CI –0.12 to –0.09; p<.001); Homeless (–0.337; 95% CI –0.35 to –0.33; p<.001); Black Homeless (–0.372; 95% CI –0.38 to –0.36; p<.001) (**Figure 2D**).

Risk–Benefit. Effects centered near zero (range, –0.086 to 0.023); no label reached ≥0.10. Homeless was most negative (–0.086; 95% CI –0.11 to –0.06; p<.001) and High-income slightly positive (0.023; 95% CI 0.00 to 0.05; p=.058) (**Figure 2E**).

Aggregated, domain-level results are in the **Supplement Table 1, Supplement Table 2**, and **Supplement Figure 1**.

## Discussion

Early evidence suggested that language models inherit human bias across medical tasks^5–9^. We expected similar distortion in trial screening, where eligibility decisions can quietly determine who gains access to therapy. Yet, under controlled conditions, eligibility judgments proved largely stable. When the clinical content and protocol were fixed, identity alone rarely changed the outcome.

This finding contrasts with prior studies showing demographic disparities in diagnostic reasoning, triage recommendations, and note generation^5,7,8^. Omar et al. reported higher odds of urgent triage and inpatient referral for marginalized groups, and Zack et al. observed racial and gender differentials in GPT-class reasoning tasks^7,8^. In contrast, our results suggest that when models are confined to explicit trial criteria, outputs remain consistent across identities.

Bias reappeared only when prompts extended beyond formal rules. In adherence, resources, and trust, socioeconomic cues, especially homelessness and low income, lowered scores, echoing societal hierarchies. These domains invite inference about behavior and capacity, and there the models mirror the patterns they have learned.

The implication is clear: the risk lies not in eligibility logic but in its periphery. If these “soft” judgments influence recruitment or resource allocation, disparities may re-enter through operational pathways rather than inclusion criteria. The appropriate safeguard is separation: keep eligibility tethered to protocol language and treat auxiliary domains as planning variables, not filters.

Our scope differs from recent equity work in clinical trial matching and medical question answering ^10^. That study introduced sociodemographic cues into prompts and found group-dependent changes in ranking and QA performance, motivating mitigation at those stages. Here, we held protocols constant, stripped prompts of social determinants of health (SDOH) content, and examined eligibility judgments as the endpoint.

Several limitations qualify these findings. We analyzed U.S. adult Phase II–III trials from 2023–2024, which may not generalize to pediatric or international contexts. Socioeconomic descriptors were simplified proxies, and standardized vignettes cannot fully capture clinical nuance. Models were tested at single time points using default configurations, and newer iterations could behave differently.

In conclusion, bias in large language model–assisted trial screening is conditional. Within fixed criteria, models reason consistently; outside them, they echo the inequities of their data. Responsible deployment in clinical research depends on preserving that boundary so that automation strengthens fairness in trial access rather than inheriting distortion.

## Data Availability

All data analyzed in this study were obtained from publicly accessible sources (ClinicalTrials.gov) and are available online.

## Authors’ Contributions

Dr Soffer and Dr Mahmud had full access to all the data in the study and takes responsibility for the integrity of the data and the accuracy of the data analysis.

Concept and design: Soffer, Omar, Nadkarni, Klang.

Acquisition, analysis, or interpretation of data: Soffer, Omar, Klang.

Drafting of the manuscript: Soffer, Omar, Klang.

Critical revision of the manuscript for important intellectual content: All authors.

Statistical analysis: Soffer, Omar.

Supervision: Klang, Nadkarni.

## Conflict of Interest Disclosures

None reported.

## Funding/Support

This work was partly supported by the Clinical and Translational Science Awards (CTSA) grant UL1TR004419 from the National Center for Advancing Translational Sciences.

## Notes

### Competing Interest Statement

The authors have declared no competing interest.

## References

1. Gupta S, Basu A, Nievas M, et al. PRISM: Patient Records Interpretation for Semantic clinical trial Matching system using large language models. NPJ Digit Med. 2024;7(1):305. doi:10.1038/s41746-024-01274-7

2. Jin Q, Wang Z, Floudas CS, et al. Matching patients to clinical trials with large language models. Nat Commun. 2024;15(1):9074. doi:10.1038/s41467-024-53081-z

3. Wornow M, Lozano A, Dash D, Jindal J, Mahaffey KW, Shah NH. Zero-Shot Clinical Trial Patient Matching with LLMs. NEJM AI. 2025;2(1). doi:10.1056/AIcs2400360

4. Srinivasan A, Berkowitz J, Friedrich NA, Kivelson S, Tatonetti NP. Large Language Model Analysis of Reporting Quality of Randomized Clinical Trial Articles: A Systematic Review. JAMA Netw Open. 2025;8(8):e2529418. doi:10.1001/jamanetworkopen.2025.29418

5. Omar M, Sorin V, Agbareia R, et al. Evaluating and addressing demographic disparities in medical large language models: a systematic review. Int J Equity Health. 2025;24(1):57. doi:10.1186/s12939-025-02419-0

6. Levartovsky A, Omar M, Nadkarni GN, Kopylov U, Klang E. Sociodemographic Bias in Large Language Model–Assisted Gastroenterology. JAMA Netw Open. 2025;8(9):e2532692. doi:10.1001/jamanetworkopen.2025.32692

7. Omar M, Soffer S, Agbareia R, et al. Sociodemographic biases in medical decision making by large language models. Nat Med. 2025;31(6):1873–1881. doi:10.1038/s41591-025-03626-6

8. Zack T, Lehman E, Suzgun M, et al. Assessing the potential of GPT-4 to perpetuate racial and gender biases in health care: a model evaluation study. Lancet Digit Health. 2024;6(1):e12–e22. doi:10.1016/S2589-7500(23)00225-X

9. Chin MH, Afsar-Manesh N, Bierman AS, et al. Guiding Principles to Address the Impact of Algorithm Bias on Racial and Ethnic Disparities in Health and Health Care. JAMA Netw Open. 2023;6(12):e2345050. doi:10.1001/jamanetworkopen.2023.45050

10. Ji Y, Ma W, Sivarajkumar S, et al. Mitigating the risk of health inequity exacerbated by large language models. NPJ Digit Med. 2025;8(1):246. doi:10.1038/s41746-025-01576-4

